# Clinician Perceptions of Generative Artificial Intelligence Tools and Clinical Workflows: Potential Uses, Motivations for Adoption, and Sentiments on Impact

**DOI:** 10.1101/2024.07.29.24311177

**Authors:** Elise L Ruan, Aziz Alkattan, Noemie Elhadad, Sarah C Rossetti

**Affiliations:** Department of Biomedical Informatics, Columbia University, New York, NY, USA; Department of Medicine, NewYork-Presbyterian/Columbia University Irving Medical Center, New York, NY, USA; Department of Surgery, NewYork-Presbyterian/Columbia University Irving Medical Center, New York, NY, USA; School of Nursing, Columbia University, New York, NY, USA

## Abstract

Successful integration of Generative Artificial Intelligence (AI) into healthcare requires understanding of health professionals’ perspectives, ideally through data-driven approaches. In this study, we use a semi-structured survey and mixed methods analyses to explore clinicians’ perceptions on the utility of generative AI for all types of clinical tasks, familiarity and competency with generative AI tools, and sentiments regarding the potential impact of generative AI on healthcare. Analysis of 116 clinician responses found differing perceptions regarding the usefulness of generative AI across clinical workflows, with information gathering from external sources rated highest and communication rated lowest. Clinician-generated prompt suggestions focused most often on clinician decision making and were of mixed quality, with participants more familiar with generative AI suggesting more high-quality prompts. Sentiments regarding the impact of generative AI varied, particularly regarding trustworthiness and impact on bias. Thematic analysis of open-ended comments highlighted concerns about patient care and the role of clinicians.

## Introduction

Recent advancements in Generative Artificial Intelligence (AI) hold significant promise for transforming healthcare through streamlining clinical workflows such as documentation, supporting empathetic and personalized patient care, and improving access to accurate medical information^1-3^. There have been examples of generative AI models performing as well as or even better than clinicians in tasks like answering patient questions^4-6^ and generating or transforming patient summaries^7-9^. However, there have also been numerous concerns cited by health professionals including the risk of bias, lack of transparency, potential inaccuracies in the form of “hallucinations” or confabulations and privacy risks^2,10-13^.

Furthermore, integration of generative AI into healthcare is a complex endeavor, requiring stakeholders such as researchers, vendors, health systems, and frontline clinicians to make strategic choices amidst time, resource, and expertise constraints. Prior studies have focused on the use of a generative AI tool to address a specific clinical task, such as answering patient questions or generating discharge summaries, but there has been limited exploration into systematically explore clinicians’ views on which specific types of clinical tasks would be most suitable for a generative AI intervention^14^. Additionally, given that the performance of many generative AI models rely on prompt quality, if clinicians are expected to design these inputs, there is a need to assess clinicians’ understanding and ability to generate prompts that would yield useful, accurate and relevant outputs^15^.

In this study, we 1) develop and use a comprehensive framework that categorizes types of clinical work, based on prior literature, to explore what types of clinical work are perceived by clinicians to have the most potential use for a generative AI tool, 2) assess the quality and type of clinical task addressed within clinician-generated prompt suggestions for a generative AI tool, and 3) examine the distribution of sentiments regarding the impact of Generative AI on healthcare on divisive stances found from prior publications, including commentary and viewpoint articles, using both structured ratings and qualitative comments.

## Methods

We conducted a cross-sectional survey targeting clinical staff employed at Columbia University Irving Medical Center. Participants were recruited from December 15, 2023 to February 15, 2024 by email or word of mouth. We relied on clinical department chairs and nursing leadership collaboration to disseminate the survey using email listservs to members of their department. All patient-facing clinical staff (e.g., attending physicians, nurse practitioners, physician assistants, registered nurses) were eligible for the survey aside from resident physicians and house staff physicians, due to a separate ongoing effort in conjunction with the Graduate Medical Education (GME) department. The survey was administered through Qualtrics, and participants were informed of the anonymous and voluntary nature of the study using an information sheet at the beginning of the survey. The study and survey instrument were approved by the Columbia University Institutional Review Board.

### Survey Design

The 32-item instrument was designed to evaluate four areas of interest: 1) Level of Familiarity with Generative AI, 2) Usefulness of Generative AI in Clinical Workflows, 3) Characteristics and Motivations for Adoption, and 4) Sentiments about the Impact of Generative AI on clinical practice

#### 1) Level of Familiarity with Generative AI

Respondents were asked to self-report their level of familiarity with generative AI tools using 3 possible categories:

Not familiar (never heard of it or have only heard of the term), 2) Moderately familiar (familiar with specific use cases or have used a generative AI tool at least once), and 3) Extremely familiar (have used a generative AI tool repeatedly and have thought about its potential applications).

#### 2) Usefulness of Generative AI in Clinical Workflows

We developed a framework (Table 1) to comprehensively categorize the types of clinical work clinicians perform using prior studies on how physicians spend their time at work, using ecological momentary assessments (EMA)^16^, time-motion studies^17^ and semi structured interviews^18^.

**Table 1.** Framework for Clinical Work Categorization.

| Category | Description | Subcategories |
| --- | --- | --- |
| **Information Gathering** | Collection of information that is necessary for delivering patient care, such as patient information, medical domain knowledge, institutional/organizational knowledge, from various sources. | **Patient/Family:** interview or physical exam |
|  |  | **Medical Record:** chart review, monitoring |
|  |  | **External Sources:** clinical resources, institutional guidelines/logistics |
| **Clinical Decision Making** | Continuous process in which gathered information is evaluated and interpreted to formulate hypotheses, diagnoses, and management plans | **Synthesis of information:** summaries, calculations/scores |
|  |  | **Clinical reasoning:** generating differential diagnoses and treatment plans |
| **EHR Documentation and Entry** | Entry of information and data into the EHR for the purposes of executing the care plan and maintaining accurate and useful records. | **Documentation/Data Entry:** structured (vital signs, diagnosis codes), unstructured (notes, social history) |
|  |  | **Order entry:** entering and sending treatment instructions |
| **Care Coordination and Provision** | Execution of the care plan through communication with patient and team members and performance of necessary tests and treatments. | **Communication:** with patients, other clinical staff, other healthcare providers regarding care plan |
|  |  | **Treatment/Management:** such as medication administration, test collection, procedures |
| **Administrative** | Processes, procedures, and requirements which are required that affect the provision of medical care services. | **Administrative:** insurance, billing, forms, compliance/regulations, scheduling |

The initial framework underwent review and refinement by the research team, which included two practicing physicians, an internist and a surgeon, and 2 clinical informatics researchers, one with prior critical care nursing experience. Using the proposed framework, the survey asked participants how useful a generative AI tool would be for each sub-category of clinical work using a 5-point Likert scale (i.e., not at all useful to extremely useful). We also included 1 open-ended question, asking participants to “suggest at least one prompt [they] would ask a generative AI tool in order to assist [them] with a specific clinical task.”

#### 3) Characteristics and Motivations for Adoption

Participants were asked to rank different factors in terms of importance when considering incorporating a new generative AI tool into clinical practice. The factors were: patient benefit, workflow, compatibility, complexity, risk, and financial factors based on review of prior surveys administered to physicians regarding adoptions of health technologies, including a national survey by the American Medical Association^19^ and the Physician-Motivation-Adoption (PMA) scale^20^.

#### 4) Sentiments about the Impact of Generative AI

We asked participants to indicate their position on nine divisive areas regarding the generative AI tools, which were selected based on published articles, viewpoints, and commentary by clinicians^1,2,5,10-13,21,22^. Participants were asked to consider the impact of generative AI on 1) time, 2) cognitive burden, 3) safety, 4) autonomy, 5) trust, 6) equity, 7) personalization, 8) standardization, and 9) the degree of impact, for both their own clinical practice and healthcare in general. We also collected any additional free-text comments by participants.

### Data Analysis

Descriptive statistics were calculated for participant demographics, including clinical role, specialty, years of practice, practice setting. Ranking and Likert scale results were tabulated and listed descriptively. Two members of the research team performed consensus coding of suggested prompts for which sub-category of the clinical work categorization framework best encompassed the clinical task the prompt was aiming to address. The same reviewers also evaluated the quality of the suggested prompts using a 5 pt scale developed by the team (Table 2). The distribution of quality of prompts was then compared between participants who self-reported different levels of familiarity with generative AI tools. Reviewers evaluating prompt quality were blinded to participants self-reported levels of familiarity with generative AI tools. Analysis of Variance (ANOVA) was used in a sub-analysis to evaluate if there was a difference in prompt quality between participants who indicated different levels of familiarity with generative AI tools. A p-value of 0.05 was selected for statistical significance. Statistical analysis was done using Stata.

**Table 2:** Scoring scale for prompt quality.

| Score | Rating | Description |
| --- | --- | --- |
| 1 | Absent | Not a request or prompt |
| 2 | Poor | Not an appropriate request for generative AI |
| 3 | Good | Appropriate request for generative AI but not phrased as a prompt |
| 4 | Great | Appropriate request and phrased as prompt but could be enhanced |
| 5 | Excellent | Well-designed and specific prompt, suggesting prior experience with a generative AI tool |

Inductive thematic analysis was performed by a single reviewer on all open-ended comments entered in the survey with generation of themes. The larger research team reviewed generated themes and codes and developed final set of themes iteratively and two reviewers performed consensus coding with the finalized list of themes.

## Results

The survey instrument was first piloted with a small group of physicians and nurses for feedback on content and readability. Minor adjustments were made based on the 24 responses to finalize the survey. After dissemination, we received 116 responses. All participants completed the survey, with 81 (70%) answering all questions including demographic questions. The average number of missed questions was 0.8. The median time spent was 7.8 minutes (IQR 4.8-11.5). Descriptive analysis of each question was performed using only participants who responded to the question. Consistent with other surveys that utilize clinical department and nursing leadership email listservs for distribution^23^, we were unable to accurately assess how many received the survey to calculate response rate. Respondents were 50% female, 54% physicians, 63% with more than 10 years since graduation, and 58% practicing in an ambulatory setting (Table 3).

**Table 3:**
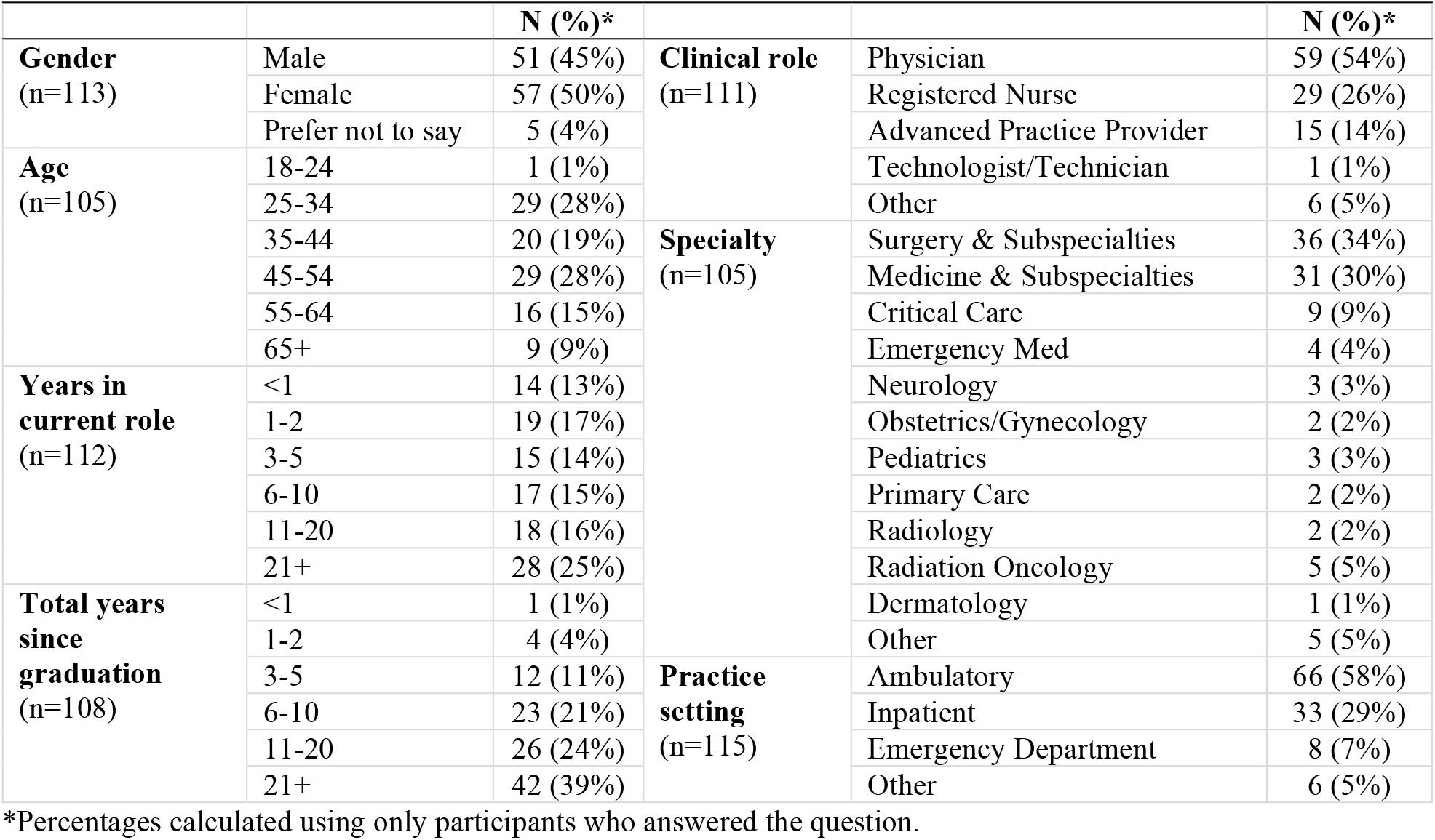
Participant Demographics.

### 1) Level of Familiarity with Generative AI

34 (29%) respondents self-reported that they were not familiar with generative AI tools, 60 (52%) were moderately familiar, and 22 (19%) were extremely familiar.

### 2) Usefulness of Generative AI in Clinical Workflows

We calculated the descriptive statistics for the ratings of potential usefulness of generative AI for subcategories of clinical work (Table 4).

**Table 4.** Comparison of respondents’ ratings for potential usefulness of generative AI tools and number of prompt suggestions for sub-categories of clinical tasks.

| Category | Subcategory | Mean Score* (SD) | Number of Prompts** |
| --- | --- | --- | --- |
| Information Gathering | Patient | 3.24 (1.24) | 2 |
|  | Medical Record | 3.82 (1.09) | 16 |
|  | External Sources | 3.90 (1.10) | 23 |
| Clinical Decision Making | Synthesis of Information | 3.62 (1.12) | 25 |
|  | Clinical Reasoning | 3.36 (1.14) | 27 |
| Electronic Health Record Entry | Documentation | 3.83 (1.07) | 14 |
|  | Orders | 3.65 (1.04) | 6 |
| Care Coordination & Provision | Communication | 3.33 (1.17) | 19 |
|  | Treatment | 3.27 (1.15) | 3 |
| Administrative | Administrative | 3.78 (1.19) | 8 |

\*Based on Likert Scale where 1 = Not at all Useful, 2 = Slightly Useful, 3 = Moderately Useful, 4 = Very Useful, and 5 = Extremely Useful \*\*Prompts coded to multiple sub-categories were included in each sub-category's count

The types of clinical work that had the highest average rating for perceived potential utility from a generative AI tool were 1) information gathering from external sources, 2) EHR documentation and data entry, and 3) information gathering from the medical record. The types of clinical work that had the lowest average rating for perceived potential utility were 1) information gathering from the patient or family, 2) treatment or management, and 3) communication.

78 responding participants submitted a total of 125 prompt suggestions. All prompts were coded to at least one subcategory of clinical work and some prompts were coded to multiple subcategories (Table 4). The subcategories with the most suggested prompts were 1) clinical reasoning (n=27), 2) synthesis of information (n=25), and 3) information gathering from external sources (n=23). The fourth most suggested with 19 prompts was communication which, per the rankings, was perceived to be one of subcategories with the least potential for use.

When coded for quality, most prompts were coded as appropriate but either not phrased correctly (44%) or could be improved (35%) (Table 5). There was a higher proportion of appropriately phrased prompts (Good and Excellent) in the extremely familiar and moderately familiar groups (50%, and 38% respectively) compared to the not familiar group (26%) although ANOVA analysis conducted to examine differences between the three groups of reported familiarity with generative AI tools did not reach statistical significance (p=0.051).

**Table 5.** Prompt quality by participant self-reported familiarity with generative AI.

| Prompt Quality | Not Familiar<br>(n=23) | Moderately Familiar<br>(n=66) | Extremely Familiar<br>(n=36) | Total<br>(n=125) |
| --- | --- | --- | --- | --- |
| **1 - Absent** | 1 (4%) | 2 (3%) | 1 (3%) | 4 (3%) |
| **2 - Poor** | 7 (30%) | 7 (11%) | 3 (8%) | 17 (14%) |
| **3 - Good** | 9 (39%) | 32 (49%) | 14 (39%) | 55 (44%) |
| **4 - Great** | 6 (26%) | 22 (33%) | 16 (44%) | 44 (35%) |
| **5 - Excellent** | 0 (0%) | 3 (5%) | 2 (6%) | 5 (4%) |

### 3) Characteristics and Motivations for Adoption

108 respondents completed the ranking of motivations for adoption. We calculated the mean ranking for each potential factor. Reimbursement and malpractice insurance coverage were ranked most important and workflow efficiency was ranked least important (Table 6).

**Table 6:** Respondents’ ranking of potential motivating factors for generative AI adoption, by importance.

| Category | Motivation | Mean Rank<br>(SD) |
| --- | --- | --- |
| Financial | 1. Can be reimbursed for time spent using it | 8.91 (1.75) |
| Risk | 2. Is covered by standard malpractice insurance | 8.56 (2.64) |
| Patient Benefit | 3. Increases patient engagement with care | 7.63 (3.03) |
| Patient Benefit | 4. Improves patient-physician relationship | 7.31 (2.65) |
| Compatibility | 5. Is consistent with my existing workflow (requires minimal or no additional steps) | 6.31 (2.69) |
| Workflow | 6. Helps to reduce stress and burnout | 5.72 (2.82) |
| Complexity | 7. Is intuitive (requires minimal or no special training) and explainable (easy to understand and use results) | 5.43 (2.49) |
| Risk | 8. Complies with data privacy and security | 5.05 (2.95) |
| Compatibility | 9. Is well integrated with my EHR | 4.62 (1.99) |
| Patient Benefit | 10. Is proven to be as good or superior to traditional care | 4.27 (2.72) |
| Workflow | 11. Improves work efficiency | 2.20 (1.68) |

### 4) Sentiments about the Impact of Generative AI

Distribution of respondents’ reported stances varied between the nine areas of focus (Figure 1). Forty-two percent of respondents agreed that generative AI will significantly change the practice of medicine for their own practice and 52% for healthcare in general. There was more division among respondents on the trustworthiness of generative AI and its impact on bias with “Neutral” being the most selected stance (26-34%) and similar numbers of respondents selecting stances on either side of the scale.

**Figure 1.**
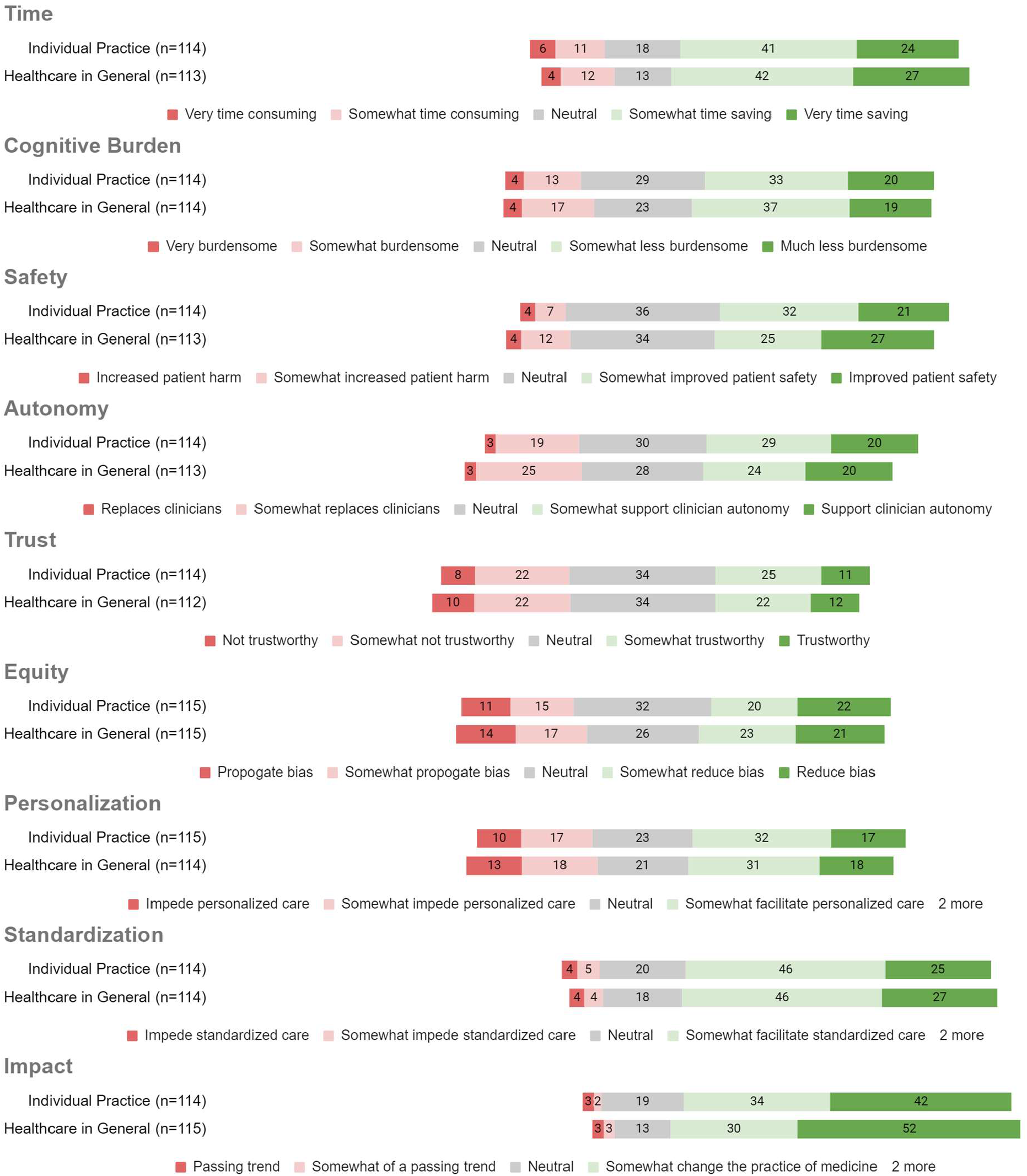
Percent of respondents reporting stances regarding impact of Generative AI

We received open-ended comments about generative AI from 40 participants. Eight themes were identified through thematic analysis (Table 7). The two most referenced themes were the *Patient care impact and concerns* and the *Impact and role of clinicians*.

**Table 7.**
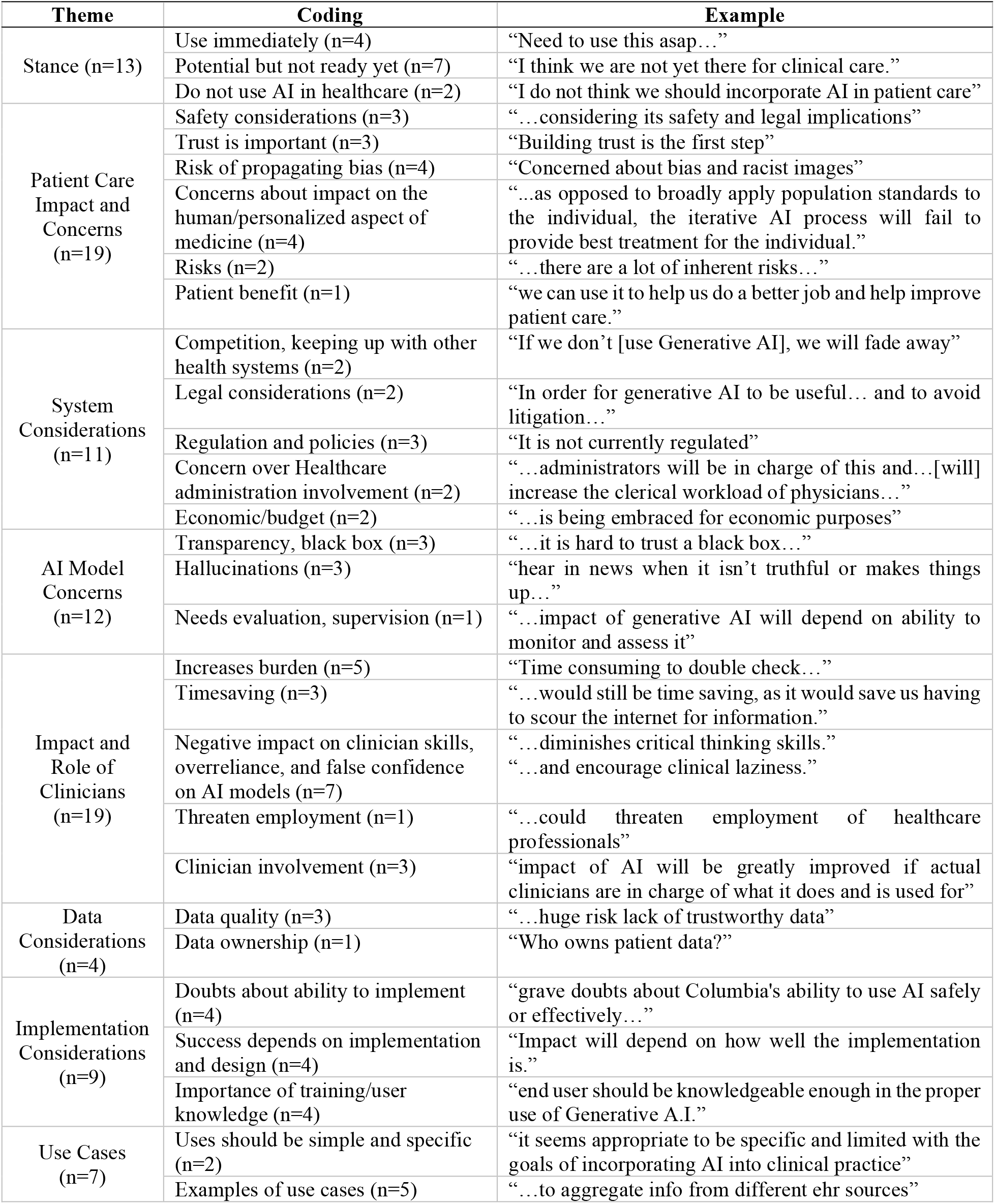
Summary of narrative comments about generative AI and healthcare.

## Discussion

This study has illuminated the considerable diversity among clinicians’ perspectives on Generative AI related to potential use across different clinical workflows. This exploratory analysis has allowed for a better understanding of how self-reported familiarity of Generative AI and clinicians ability to create an appropriate prompt example may be important to consider when evaluating clinician rankings of potential uses of Generative AI. We see this work as an initial exploration to guide clinically relevant areas to focus Generative AI innovations while still recognizing the large gaps in the “state of understanding” of Generative AI by clinicians that could potentially bias these rankings.

Participants’ views on the usefulness of generative AI tools for various clinical tasks, as well as the types of tasks they sought to address through their prompt suggestions, were not entirely consistent. While information gathering of external sources was perceived to be one of the most useful tasks and received many suggested prompts, the clinical tasks with the most prompt suggestions were in the clinical decision making category as well as communication, which was actually perceived to be one of the least useful tasks by participants. The differences in prompt submission and perceived usefulness may also be due participants submitting prompts based on ease of generation rather than targeting the most useful tasks. Especially given the variability in familiarity with AI tools, participants may have had difficulty constructing prompts for their desired applications. Furthermore, to be inclusive of many different types of clinical workflows, the subcategories of the work were still general, and the inconsistencies may have been due to differing perceptions about tasks within each subcategory, such as communication with patients versus communication with other clinicians. These differences indicate that assessing clinicians’ perceptions of emerging technologies may benefit from collecting both quantitative and qualitative data by observing clinicians engaging in a proposed process in addition to theorizing about their potential experience.

Analysis of prompt suggestions revealed a broad spectrum of quality, with participants who were more familiar with generative AI tools overall submitting higher-quality prompts. Many studies have evaluated clinicians’ perceptions and experience regarding health technologies, but an important consideration is their familiarity and proficiency with the technology. This is especially relevant for generative AI as performance of the tool is strongly linked to the quality of the prompt^3,15^. This sentiment was captured in the open-ended comments where the importance of end-user knowledge and training in the success of a generative AI tool was emphasized.

Our method of having clinicians rank their position on nine dichotomous stances regarding the impact of generative AI helped gather a more comprehensive high-level view on the overall sentiments of a large heterogeneous group. We could see where perspectives converged, such as generative AI “will significantly change the practice of medicine” and “will somewhat facilitate standardized care” and where there was more division, such as generative AI’s impact on clinician autonomy and trustworthiness.

We supplemented these general sentiments with an opportunity for participants to express more detailed perspectives through free text comments and several salient points arose both within and outside our nine areas of focus. In every theme there was variation in perspectives. While some clinicians inquired about the implementation of generative AI tools as soon as possible, others expressed that AI should not be used in healthcare in any capacity. There were many concerns regarding impact on patient care, such as safety, bias, impeding personalized care and regarding the models themselves, particularly the lack of transparency and potential for hallucinations (confabulations). A recurring theme, *Impact and role of clinicians*, included varied opinions on whether generative AI tools would reduce burden or save time. There was also concern that, in addition to overreliance and false confidence in these models, clinicians may become less skilled in their clinical decision making. Respondents also mentioned the importance of having clinicians, not just hospital administrators and regulators, involved to avoid a misalignment of goals and subsequently an increase in clinician documentation burden and workload.

Limitations of our study mostly relate to sampling; we used convenience sampling through professional email listservs at our organization and, therefore, were unable to assess the response rate, consistent with other survey studies of clinicians. There was additionally a small amount of missing data for certain questions, most notably for demographic questions. For the survey categories, the ranking of motivations for adoptions had the most amount of missing data with 8 respondents (7%) that did not answer the question. It is possible the participants who did not answer the question represent a group that is less motivated to adopt use of a generative AI tool. Our sample, though targeted broadly, was also strongly skewed towards physicians and nurses, and all respondents were employees at a single tertiary academic medical center in an urban setting. Additional work should be done in examining differences in responses by clinical role, specialty, practice setting as well as expanding the target pool of respondents to include clinicians from other health systems and settings.

Despite the variation in perceptions regarding the use and impact of generative AI tools in healthcare, clinicians generally believe that the use of such tools is inevitable and will significantly impact the practice of medicine over time. Respondents emphasized the need for clinician involvement to guide the development and implementation of tools to ensure that the appropriate types of work are being addressed and that clinician skills are not being negatively impacted. Especially with Generative AI, training and proficiency of clinicians in using these tools should be a focus of implementation.

Our findings demonstrate variability not only in clinician perceptions, motivations and sentiments regarding the use of generative AI tools for clinical care but also that findings may differ depending on the method of evaluation and clinician understanding and experience with such tools. Future studies should explore the relationship between quantitative scoring of clinician perceptions and qualitative assessment of real-world behavior as well as how clinician proficiency and experience with a technology may significantly influence efficacy and impact of technologies like generative AI, where prompt engineering plays a pivotal role.

## Conclusion

Our study used a semi-structured survey to explore clinicians’ perspectives on the potential utility and impact of generative AI. Our mixed-methods analysis allowed for analysis of clinician rankings and other self-reported data as well as an understanding the sentiments of a diverse and heterogeneous group. We not only observed differing perceptions regarding the utility of generative AI tools for diverse clinical tasks, but we also captured varying sentiments regarding the different potential impacts on healthcare. Clinicians’ self-reported familiarity and experience with generative AI emerged as an important factor to consider when engaging clinicians in Generative AI use case identification and development processes for addressing the unique challenges of different clinical workflows.

## Data Availability

All data produced in the present study are available upon reasonable request to the authors

## Acknowledgements

The primary author acknowledges the support of the NewYork-Presbyterian Information Technology through the Clinical Informatics Fellowship which provides funding for postdoctoral training and education for physicians.

